# Is this the beginning or the end of COVID-19 outbreak in India? A data driven mathematical model-based analysis

**DOI:** 10.1101/2020.04.27.20081422

**Authors:** Anupam Singh, Jhilik Dey, Shivam Bhardwaj

## Abstract

India has experienced an early and harshest lockdown from 25^th^ March 2020 in response to the outbreak. However, an accurate estimation of the progression of the spread of infection and the level of preparedness to combat this disease are urgently needed. Using a data-based mathematical model, our study has made predictions on the number of cases that are expected to rise in India till 14^th^ June 2020. The epidemiological data of daily cases have been utilized from 25^th^ March (i.e., the first day of lockdown) to 23^rd^ April 2020. In the study, we have stimulated two possible scenarios (optimistic and pessimistic) for the prediction. As per the optimistic approach of modelling, COVID-19 may end in the first week of June 2020 with a total of 77,900 infected cases including 2,442 fatalities. However, the results under the pessimistic scenario are a bit scary as it shows that a total of 283,300 infected cases with 10,180 fatalities till 14^th^ June. To win the battle, 10 weeks of complete lockdown is much needed at least in the infected states and the union territories of India. Alternatively, the isolation of clusters (hotspot regions) is required if India wants a resume of some essential activities.

## Introduction

The sudden outbreak of COVID-19 which has originated in Wuhan (China) has engulfed almost the whole world. On 11 March 2020, World Health Organization (WHO) declared novel coronavirus disease (COVID-19) outbreak as a pandemic and urged all countries to take necessary actions for treating, identifying active cases, and preventing transmission to save people’s lives. The worldwide spread has created panic amongst the people and hence it has become a challenge for governments to control and mitigate COVID-19 as soon as possible. As of 3^rd^ May 2020 (2 May 2020, 05:30; GMT+5:30), there have been 3,272,202 confirmed cases with 230,104 deaths (WHO)

Coronavirus is one of the families of various viruses that are responsible for respiratory diseases like cough and cold, severe acute respiratory syndrome (SARS), and Middle East Respiratory Syndrome (MERS). A zoonotic coronavirus (CoVs), provisionally known as 2019 novel betacoronavirus (2019-nCoV) or the severe acute respiratory syndrome coronavirus 2 (SARS-CoV-2) has been indentified first in the Wuhan city of China’s Hubei Province, and it is now rapidly spreading to the rest of the world [1]. As in late December 2019, at Wuhan, some patients were admitted to the hospital and were diagnosed with severe pneumonia of unknown sets of reasons associated with clinical symptoms of high fever, dry cough, and dyspnea. Many of those clusters of people were linked to the Huanan seafood and wet animal market there [2]. Following activating surveillance and further etiologic and epidemiologic investigations of their respiratory samples, China informed this outbreak to the WHO on 31^st^ December 2019 [3]. From the throat swab samples of the affected patients, the causative agent was identified by the Chinese Centre for Disease Control and Prevention (CCDC) on 7^th^ January 2020 and the virus was named as Severe Acute Respiratory Syndrome Coronavirus 2(SARS-CoV-2) by the International Committee on Taxonomy of Viruses (ICTV). Thenceforth, the disease was named as Coronavirus Disease 2019 (COVID-19) by WHO on 11^th^ February 2020 (World Health Organization, WHO Director-General’s Remarks at the Media briefing on 2019-nCoV on 11 February 2020; https://www.who.int/dg/speeches/detail/who-director-general-s-remarks-at-the-media-briefing-on-2019-ncov-on-11-february-2020).

### The virus (SARS-Co V-2)

CoVs are large enveloped positively single-stranded RNA viruses that cause respiratory illness in animals as well as humans ranging from a mild cold to life-threatening pneumonia. In the coronaviridae family, seven contagious virus affects human, four are endemic respiratory virus responsible for causing 15–30% of common cold infection(mild respiratory illness), two of them i.e. SARS-CoV-1 and MERS are considered to be epidemic while SARS-CoV-2 is now has been considered as pandemic [4]. Studies from genetic sequencing of SARS-CoV-2 have shown 50% similarity with MERS-CoV and more than 80% similarity with SARS-CoV [5]. Due to a high level of genomic similarity it can be suggested that both SARS-CoV and SARS-CoV-2 have originated from the bat which acts as their natural primary host [6].

### The disease (COVID-19)

The third CoV outbreak in the last two decades is the COVID-19 where a novel CoV (nCoV) has appeared in the human population by crossing species after Severe Acute Respiratory Syndrome CoV(SARS-CoV-1) outbreak in south China, 2002 and the Middle East Respiratory Syndrome CoV(MERS-CoV) outbreak in Saudi Arabia, 2012 [7]. The transmission of infection occurs through airborne droplets produced during coughing and sneezing by the affected person as well as asymptomatic persons both before and after the arrival of symptoms [8]. The incubation period of the virus ranges from 2–14 days which is a quite long duration available for the transmission of infection. Clinical symptoms associated with COVID-19 are generally sore throat, cough, fever, fatigue, and dyspnea. Sometimes, in some patients (often persons with comorbidities and elderly people), following these aforesaid symptoms after nearly 7 to 10 days, the disease may progress to Acute Respiratory Distress Syndrome(ARDS), pneumonia, respiratory collapse and multiple organ failure leading to death[3]. Hence, some patients experience a mild disease while in others it may progress to a lethal situation while some people always remain asymptomatic. The case fatality rate of SARS-CoV-1 is nearly 9.6% while that of MERS-CoV is around 34% [7]. But for the COVID-19 fatality rate of the cases is approximately ranged between 2–3% which is less than that of the previous two outbreaks i.e. SARS-CoV and MERS-CoV while it has the highest transmission competence with less pathogenesis [3].

## COVID-19 outbreak in India

India started the fight when the government evacuated 767 people through airlifting. Foreign travelers were the main reason behind the spread of viruses. The first case of COVID-19 in India was detected on 30^th^ January 2020. A student who returned from Wuhan city was found to be coronavirus positive. Another patient who returned from Italy was found to be infected with this virus. Later on few Italian tourists and their guide were detected positive in Rajasthan. Since then there occurred a gradual rise in the number of cases in all regions of India. Initially New Delhi and Maharashtra were the states with the highest rate of infection. As of 2^nd^ May 2020 (08:00 GMT+5:30), according to the Ministry of Health & Family Welfare (MoHFW), the total number of confirmed cases reported in India is 39,980 with 1301 deceased and 10,632 recovered. (www.mohfw.gov.in/) India is a highly dense country of 1.35 billion populations with limitations in the proper health care system and infrastructure. Hence, it is at a higher potential risk of infection than the other developed countries [9]. So, India to a greater extent has to aim at non-compulsive community intervention with people’s commitment and assurance at enough lucidity [10]. These preventive and control strategies might help in the mitigation of infection by hindering person to person transmission at the community level as well as national level. To fight against COVID-19, the Indian government has taken unprecedented measures like lockdown, cancellation of visas, flights, trains, and conducting awareness campaigns which creates hope of successful control of the situation [11].

### Methods and Models

#### Exponential Model

In epidemiology, researchers use different mathematical models to estimate the outbreak. The selection of a model depends on the number of factors of that region or country such as current growth rate of disease, demography (e.g., age and population density), weather or humidity changes, immunity, healthcare facilities, and government interventions [12],[13]. Studies suggest that most pandemics follow exponential growth in their initial stages and then flatten out [14]. Different researchers have used different mathematical models to predict the spread of COVID-19. The current study assesses the suitability of each model.

The exponential model is good for estimating the outbreak in the short run of the first phase as the pandemic spread exponentially. Thus, if the number of infected cases (I) over time (t) with infection rate (β) is known, the spread can be calculated by,

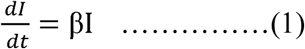

Integrating above equation 1, we get-

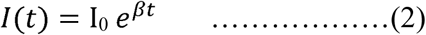

Where *I_0_* is a constant and can be obtained by fitting with the data. And, *I* is the cumulative number of cases including individuals recovered and deceased. A J-shape is formed in the exponential growth phase of a pandemic. The exponential growth model fails to predict decay (downward movement) and the plateau of the curve.

As of 19^th^ April 2020, India is in the 2^nd^ phase of disease transmission, where total cases of infections are increasing steadily. Thus, exponential growth will not be able to predict accurately.

#### Logistic Model

Unlike the exponential growth model, this model predicts the growth of the disease spread under the constraints. We mean constraints in terms of redefining the number of susceptible after the interventions imposed by the government (especially, restriction People’s mobility, strict lockdown, and dedicated medical facilities). Thus, change in the number of infected (I) over time (t) is calculated by

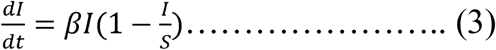

Where I is total number of accumulated infected cases, β>0 infection rate, and S is susceptible people.

If I(0)=I_0_>0 is initial number of infected cases.

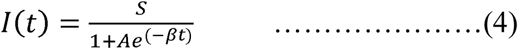

Where A= (S/I_0_)-1 and I_0_ is initial number of invectives. And S is number of susceptible people in the population.

#### SEIR Model

The widely used framework for predicting the population growth or disease spread is ‘Susceptible-Infectious-Removed’ (SIR). However, in pandemic (COVID-19 cases) like situations; the ‘Susceptible-Exposed-Infectious-Removed’ (SEIR) model has better accuracy as the model accommodates many factors which affect the growth of spread. Given the present situation in India after several interventions by the government, we formulated the model equation as follows

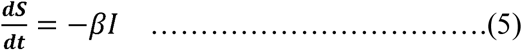

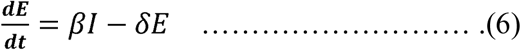

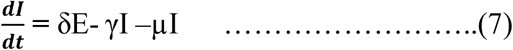

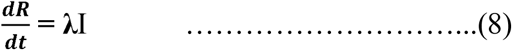

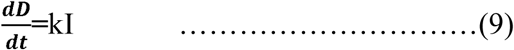

Where S is susceptible individuals; E is infected individuals but asymptomatic; I is infected and symptomatic individuals. The λ and k are the rate of recovery and death, respectively. dR and dD are cases of recoveries and deaths over time (t), respectively. We numerically simulated the SEIR model to project forward. The following parameters (Table 1) have been used in this study.

**Table 1:** Parameters of the study

| Parameter | Notation | Value | Remarks | Reference |
| --- | --- | --- | --- | --- |
| Initial population size | No | 350 million | Constant | Census data |
| Initial susceptible population | So | 0.5No | Constant | Assumed |
| Transmission rate | $\beta$ | {0.80, 1.10}/day | A stepwise function | Fitted |
| Mean latent period | $\delta$ | 3 | Constant | Wu et al. (2020) |
| Mean infectious period | $\gamma$ | 5 | Constant | Wu et al. (2020) |

**Note:**

Note
1. *Total urban population of the top 20 infected Indian states considered as initial population*.
2. *To estimate total susceptible, 0.50% of total urban population of the top 20 highly infected Indian states was taken*.

### Data

We obtained daily updates of the number of reported confirmed cases for the COVID-19 pandemic across states in India from the Ministry of Health and Family Welfare website (www.mohfw.gov.in/index.html) and COVID-19 India (www.covid19india.org/). The data was collected from the first day of lockdown (e.g., March 25, 2020) to 2^nd^ May, 2020 for our analysis. The disease progression from 4^th^ May to 14^th^ June is analysed. We projected the progression of virus spread for the next 42 days from the end of the 2^nd^ phase of lockdown (e.g., 3^rd^ May). Given the incubation period of 14 days for COVID-19, the duration was divided into three cycles of 14 days each (14*3=42 days) in our analysis.

### Scenario Analyses

The coming weeks are going to be tough for India. The government is mulling over the actions to be taken after the lockdown 3.0, which is ending on 17^th^ May. The government has the challenge of managing two contradicting things simultaneously; first one is to fight the pandemic by the virtue of isolation and social distancing, and another is to the keep wheel of the economy moving. A well thought decision will be very crucial for the people and the country as a whole. Under this circumstance, we simulated the data in two scenarios (optimistic and pessimistic). The fact under each scenario is analyzed for the predictions.

1. **A minimum of 70 days complete** lockdown with no essential activities can prove most effective to combat the COVID-19 outbreak in India. According to Richard Horton (editor in chief, The Lancet), the Indian government’s decision of lifting the lockdown will prove worse because the second wave of the disease can be more detrimental. The investment and efforts of the early lockdown will go in vain if the lockdown is withdrawn. With 10 weeks of successful lockdown in Wuhan, China proved to be effective for containing the spread.
2. **Lockdown for 70 days with resuming the essential activities** can prove effective in controlling and preventing infection spread to some extent. But after 3^rd^ May 2020, the permission to restart some essential activities may lead to the spread the disease further. There are chances that infection may spread among the workers first and then to the people who are in their contacts and the proximity. The government, in this case, must be watchful.
3. **Withdrawal of lockdown-** if India rushes to exit the lockdown, there will be a devastating circumstance. No contact tracing will be possible, and the situation will be out of control. One can’t even imagine the situation when the transmission goes into the community with limitations of per capita health care facilities in the country.

## 5 Results and Discussion

The rate of COVID-19 transmission in India is growing steadily. However, so far, India is in a better position in the battle as compared to many other countries where spread was almost equal once but now total infected cases in those countries have gone up much fold. Given the very complex demography (e.g., population density, diversity, belief system), it raised challenges to the government in imposing the sudden restrictions on people to curb the virus spread. However, the Indian government managed to take bold and timely actions such as lockdown and mobility restriction to fight against COVID-19. Thus, considering the current situation in India, projection using a linear growth model is found suitable for this study. Now that India is resuming certain economic activities, it poses further speculations if the cases may rise again due to people’s mobility and gathering. Under this circumstance, we considered two scenarios in our analysis; optimistic, and pessimistic. The optimistic scenario indicates that the spread of the virus follows the current trend with a complete lockdown in the infected states. The pessimistic scenario indicates that the growth of spread would be with a higher transmission rate due to people’s noncompliance of social distancing norms due to removal or relaxation of lockdown.

Figure 1 shows the actual daily cases. From the curve, it is evident that the daily new cases are increasing steadily. Figure 2 depicts the progression of the outbreak in both optimistic and pessimistic scenarios. The optimistic models are statically significant at p<.01 with R^2^ values of 91.8% for linear, and 85% for growth model. Under this scenario daily new cases increase and reach a peak in the second week of May. Thereafter, downward movement of the curve starts, and likely to touch the bottom on 14^th^ June with approximately 40 new cases daily. Thus, according to optimistic models, the COVID-19 pandemic in India will end in the 3rd week of June 2020 with a total of 77,900 cases.

**Figure 1:**
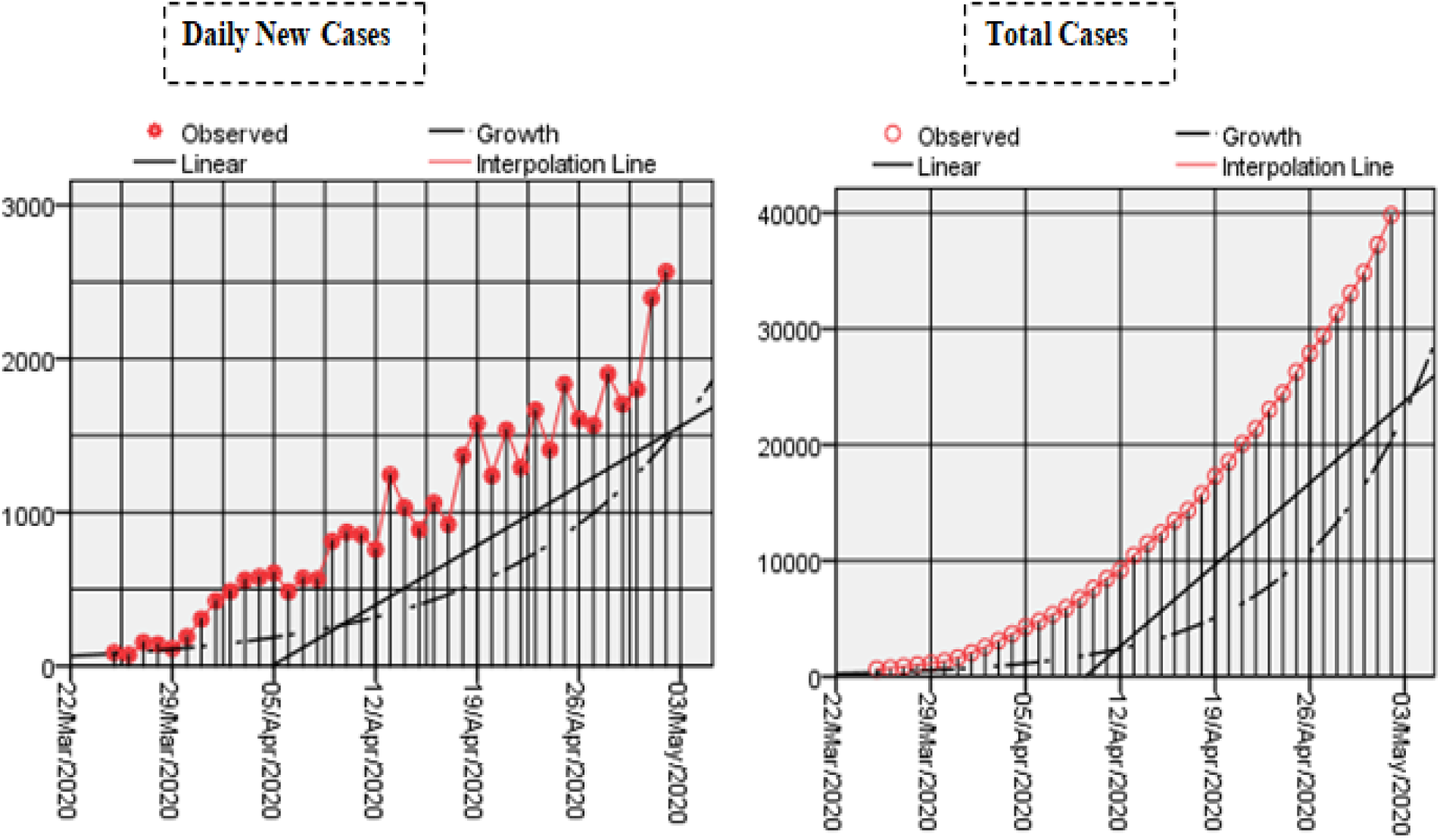
Actual daily and total cases from 25/March/2020 to 2/May/2020

**Figure 2:**
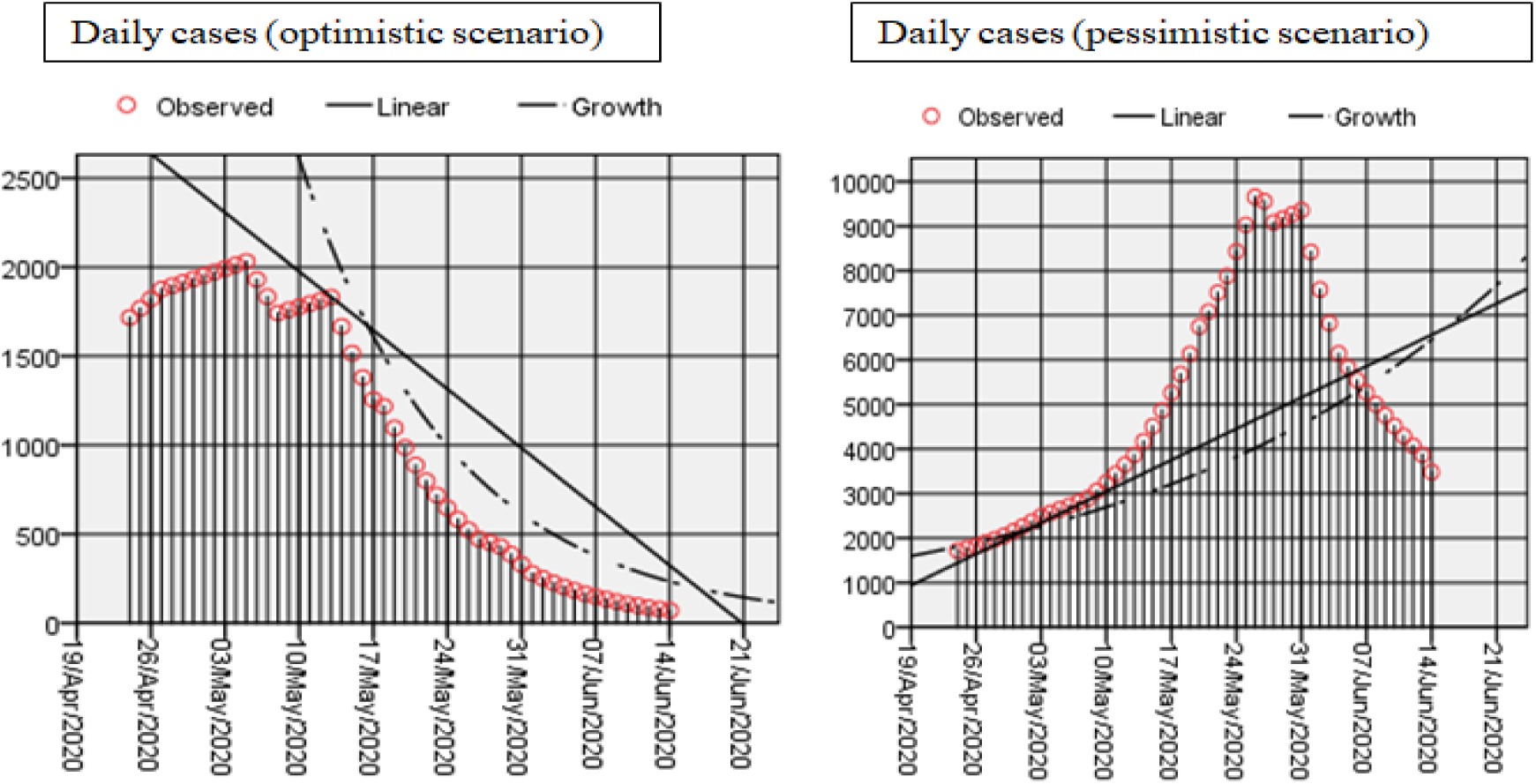
Prediction on daily new cases till 14th June 2020

The pessimistic scenario considers the worst possible cases in a certain outbreak situation. According to the results, the pessimistic models are statically significant at p<.01 with R^2^ values 49% for linear and 56% for the growth model. As per these models (linear and growth), the daily new cases will reach a peak between 24^th^ to 31^st^ May (Figure 2, pessimistic scenario) with total cases of 207,780 (Figure 4). As of 14^th^ June, 283,348 individuals will be infected with COVID-19. Under this scenario the end of the pandemic is expected to be delayed by 4–5 weeks and hence will likely to vanish in the second week of July 2020.

**Figure 3:**
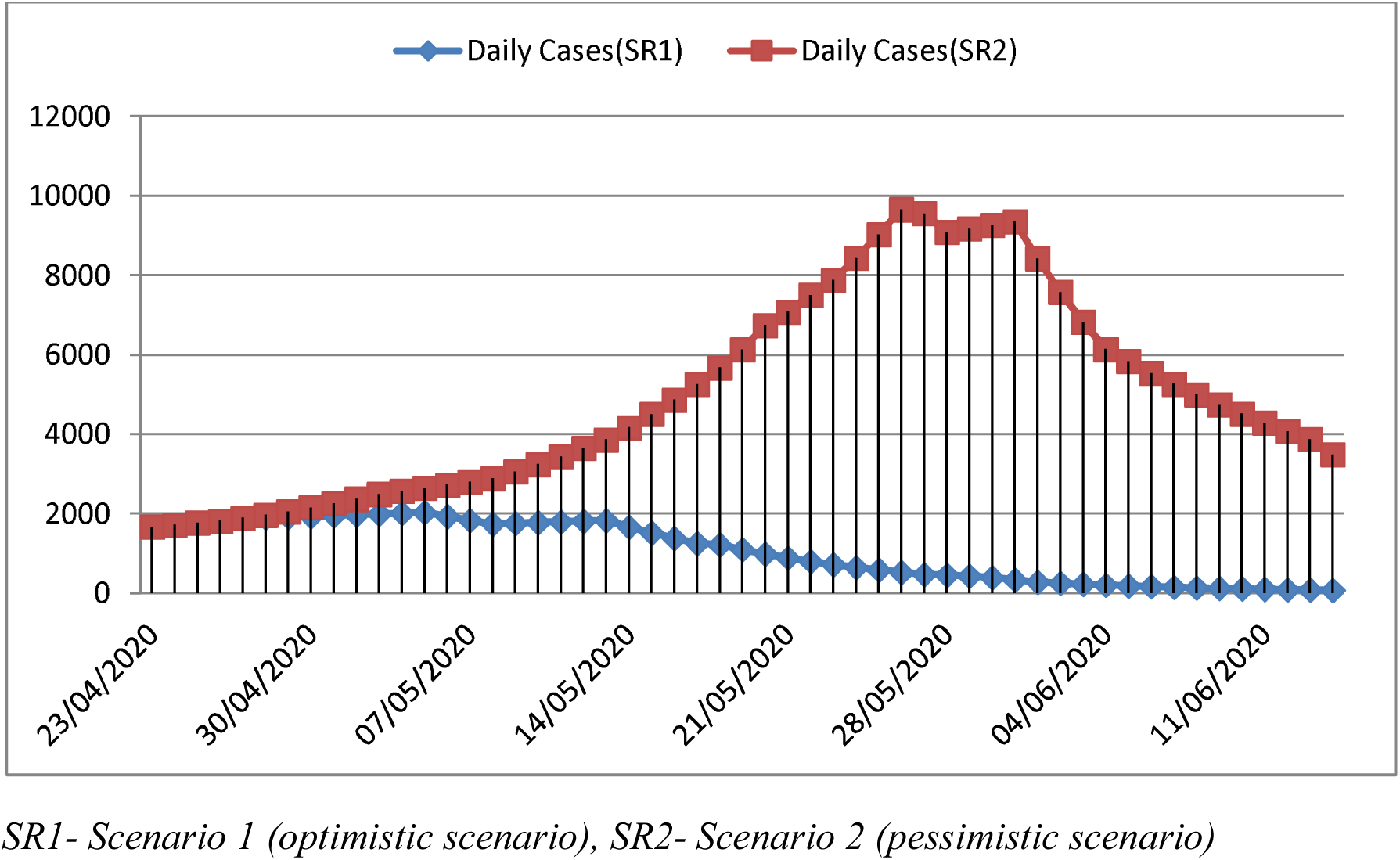
Daily new cases (Optimistic vs. Pessimistic scenario)

**Figure 4:**
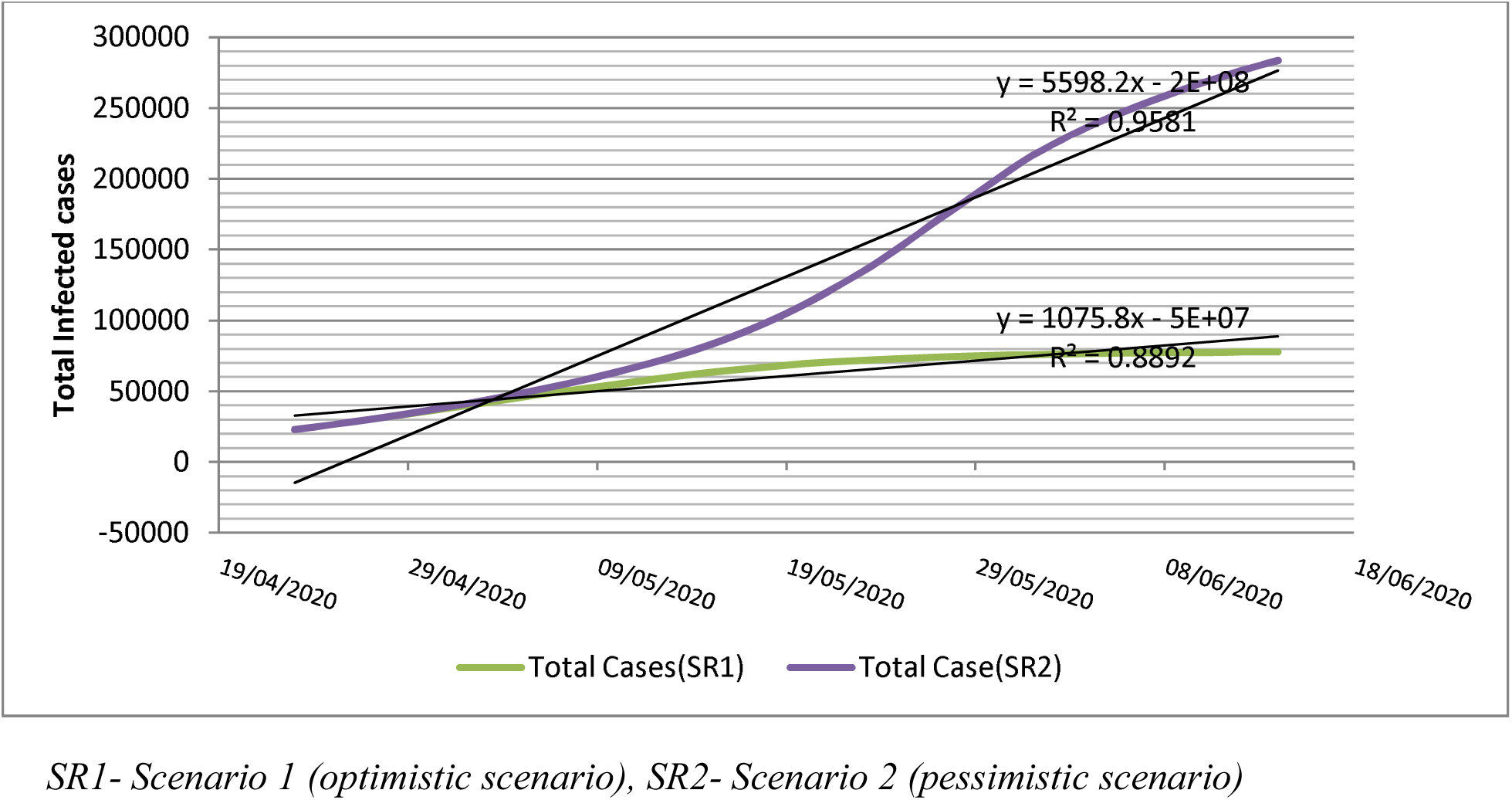
Projection of total COVID-19 Cases in India (Optimistic vs. Pessimistic scenario)

Figure 5 presents a projection of the total death toll in India till 14^th^ June 2020. As per the results obtained, in the optimistic scenario, total fatality reaches to 2,442. While in the case of a pessimistic scenario, the total fatality would go up to 10,180.

**Figure 5:**
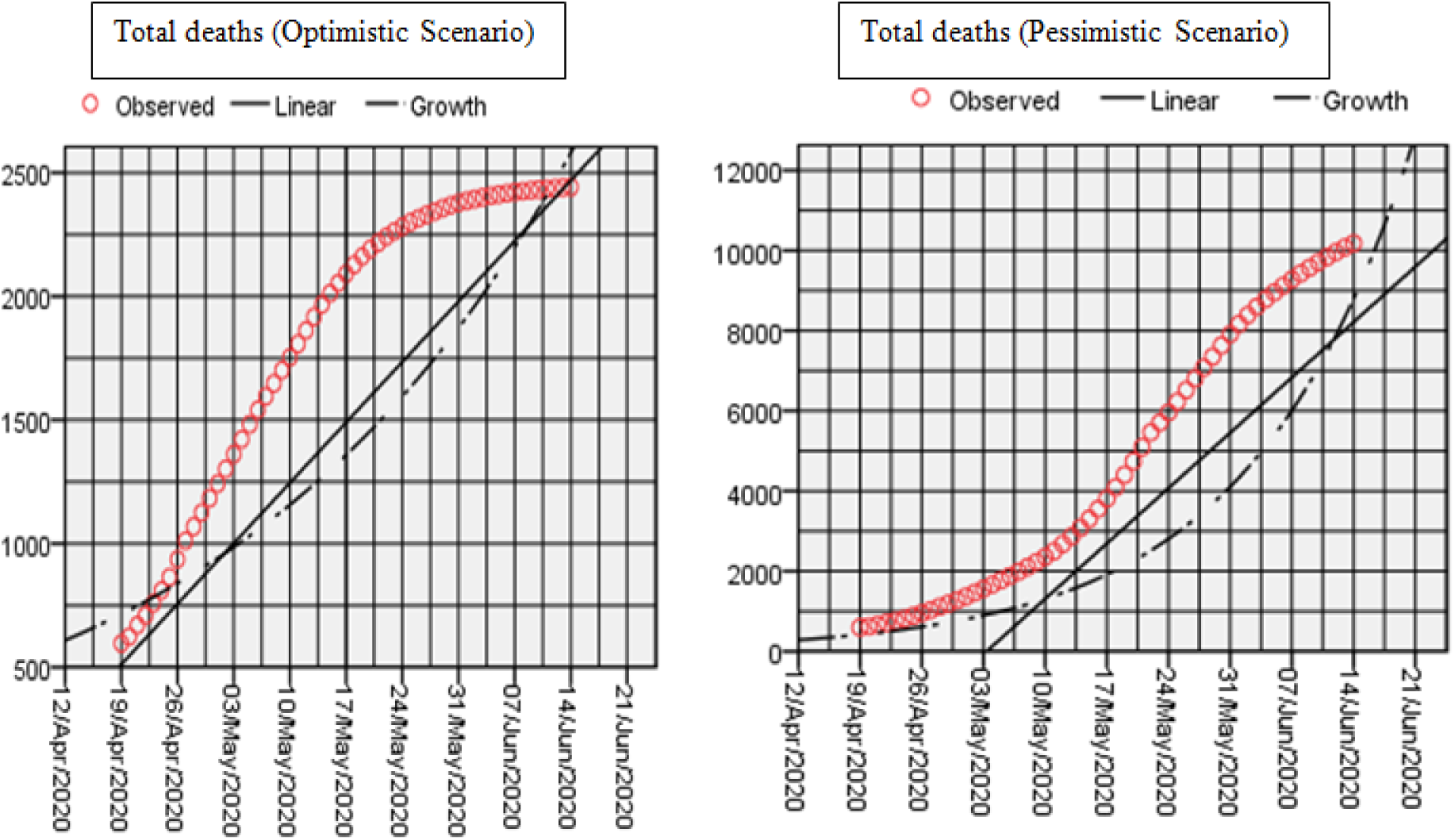
Projection of total deaths in India

## Conclusion

We have estimated the spread of COVID-19 using mathematical modelling. The data were taken from the very early days of lockdown to specifically examine the impact of lockdown in curbing the disease outbreak. In the current study, we have used different forecasting models under two scenarios, the first scenario of optimistic approach depicting the progression of infection in the presence of government interventions like social distancing, complete lockdown, public awareness, travel restrictions, and another scenario of a pessimistic approach of the infection progression without (or less) such interventions. By validating all analyses done, if India can manage interventions implemented, the daily infected cases have gradually restrained from mid-May, 2020. Hence, under an optimistic approach, the total number of cases is expected to come down from the 3rd week of May 2020. Results under optimistic conditions confirm the total 77,900 infected cases till 14^th^ June. However, the results of the pessimistic scenario estimation are horrifying. It shows that the spread will be quite high in the mid of May. The total case, till 14^th^ June will reach 283,300. Therefore, the complete lockdown (at least in the infected Indian states) and travel restrictions are much needed in the fight against a highly contagious virus like COVID-19 to avoid the pessimistic scenario in the country. It should further be noted that in both scenarios we assumed no local transmission. For a country like India with high population density and poor health care systems, it would be quite difficult to mitigate outbreak without any of such serious laws and steps enacted by both state and central government. The 10 weeks of lockdown will be required in the fight against COVID-19. Additionally, with more rigid lockdown mass screening, isolation, enforced quarantine and proper medical treatment should be executed meticulously for pandemic mitigation.

These predicting models thus provide an insight into the epidemiological status in the country by forecasting the potential propensity of a disease outbreak. Thus, on an analytical basis, it would help in the execution as well as manipulation of all current interventions and approaches to combat the current pandemic in India.

Our present model signifies the prediction capability of COVID-19 growth in India by considering the effects of interventions (both government and self) as preventive and control measures undertaken since the time of spread. But these models are devoid of the impact of certain biological factors like differential immunity, previous and present health status of person i.e. comorbidities associated and passive immunization status. The physical factors like temperature, humidity, their influence have not been considered in the modeling.

### Implications and suggestions

According to Boston Consulting Group (BCG) analysis (source, John Hopkins University), countries that implemented complete lockdown are in a better position in the combat with the COVID-19. India has also taken the early steps such as lockdowns, travel restrictions, and identifications of infected zones, etc.

If a lockdown is fully or partially withdrawn in a premature state of an outbreak, then there will be higher chances of the exponential growth of spread. Complete lockdown signifies a gradual decrease in transmission rate with time due to more contact inhibition. To date, the only effective way is to do proper mass scale random testing along with testing suspects. The aggressive contact tracing efforts are much needed to minimize the fluidity of the spread. A strict lockdown will be helpful for India in breaking the chain and contact tracing.

According to the world’s renowned virologist, Ian Lipkin, it is difficult to predict the end of the pandemic in India. However, he advised India to follow the two strategies to win the battle; first is the aggressive testing, and second is the isolation of clusters (hotspots region). If these steps are taken seriously, India may go with an option of relaxation of essential activities, and with some restrictions; people can resume their daily activities. So, contact tracing is most essential in this scenario. Discovering the contact history of the infected or suspected person is an urgent measure that has to be undertaken. The identification of the most affected areas (hotspots) and isolating them completely will be effective in further prevention. There are moderate chances of infection spread in this scenario with a capacity of control by the government to some extent.

## Data Availability

World Health Organization (WHO)

https://www.who.int/

https://www.worldometers.info/coronavirus/

## Ethical approval

Approval was not required.

